# Reconstructing subdistrict-level population denominators in Yemen after six years of armed conflict and forced displacement

**DOI:** 10.1101/2022.03.07.22272007

**Authors:** Francesco Checchi, Emilie Sabine Koum Besson

## Abstract

**Introduction:** Yemen has experienced widespread insecurity since 2014, resulting in large-scale internal displacement. In the absence of reliable vital events registration, we tried to reconstruct the evolution of Yemen’s population between June 2014 and September 2021, at subdistrict (administrative level 3) resolution, while accounting for growth and internal migration.

**Methods:** We reconstructed subdistrict-month populations starting from June 2014 WorldPop gridded estimates, as a function of assumed birth and death rates, estimated changes in population density, net internal displacement to and from the subdistrict and assumed overlap between internal displacement and WorldPop trends. Available displacement data from the Displacement Tracking Matrix (DTM) project were subjected to extensive cleaning and imputation to resolve missingness, including through machine learning models informed by predictors such as insecurity. We also modelled the evolution of displaced groups before and after assessment points. To represent parameter uncertainty, we complemented the main analysis with sensitivity scenarios.

**Results:** We estimated that Yemen’s population rose from about 26.3M to 31.1M during the seven-year analysis period, with considerable pattern differences at sub-national level. We found that some 10 to 14M Yemenis may have been internally displaced during 2015-2016, about five times United Nations estimates. By contrast, we estimated that the internally displaced population had declined to 1-2M by September 2021.

**Conclusions:** This analysis illustrates approaches to analysing the dynamics of displacement, and the application of different models and data streams to supplement incomplete ground observations. Our findings are subject to limitations related to data quality, model inaccuracy and omission of migration outside Yemen. We recommend adaptations to the DTM project to enable more robust estimation.

## 1 Background

In scenarios of crisis due to armed conflict or natural disasters, both governments and humanitarian actors require accurate population denominators to plan, mobilise resources for, implement and monitor the performance of services to the affected population [1, 2]. Most contemporary crises occur in settings with weak vital events registration and infrequent census exercises, often resulting in uncertain population figures even prior to the crisis: this uncertainty is compounded during the crisis itself by displacement within and outside the crisis region, which can disproportionately depopulate certain locations and stretch the hosting capacity of regions that receive displaced persons. Furthermore, displacement patterns can be complex, with households disintegrating and individuals experiencing multiple waves of displacement and/or return to communities of origin. The effects of the crisis on birth and death rates can also affect the trajectory of population size. While excess mortality due to armed conflict crises is extensively documented [3], there is less evidence on the effects of conflict and its downstream consequences on fertility [4]: it is plausible that insecurity and loss of livelihoods would lead families to delay births, but an inverse effect could result from reduced ability to adopt contraception.

Yemen has been affected by widespread armed conflict since late 2014. As of end 2021, some 24.1M Yemenis were in need of humanitarian assistance and 3.3M were estimated to be internally displaced persons (IDPs) [5]. Yemen does not have a functional birth and death registration system. As part of a study to estimate crisis-attributable mortality in Yemen, we wished to generate a dataset of population denominators stratified by month and subdistrict, which, in Yemen, equates to administrative level 3 (below governorate and district).

## 2 Methods

### 2.1 Study population and period

This analysis encompasses the entire country of Yemen from June 2014 to September 2021. The official ‘gazetteer’ (geographical index) published by the United Nations Office for Coordination of Humanitarian Affairs (OCHA) and Yemen’s Central Statistical Office (CSO) lists 22 governorates, 334 districts and 2149 subdistricts [6]; many cities are divided into districts only, i.e. district and subdistrict are one. Due to insufficient geographical detail in available data, our analysis omits refugees from other countries living in Yemen (141,000 as of August 2021 [7]) and assumes no migration out of Yemen (refugees or economic migrants). Yemeni refugees and asylum seekers numbered 54,000 worldwide according to the United Nations High Commissioner for Refugees as of September 2021 [8], though an uncertain number may not be registered at all. Other Yemeni emigrants were estimated at 1,268,000 in 2019, up from 1,112,000 in 2015 and 877,000 in 2010 [9].

### 2.2 Data sources

#### 2.2.1 Population estimates

Yemen conducted its last census in 2004 [10]. The UN World Population Prospects [11] provide country-wide yearly projections from this baseline, reflecting assumed natural growth. The WorldPop project redistributes these projections across space, with 100 m^2^ resolution, by applying a geospatial model that predicts population density using a variety of remotely sensed climate, topography, illumination, transport network, urbanisation and other land use variables (see https://www.worldpop.org/methods, Stevens et al. [12] and Linard et al. [13]). WorldPop annual estimates were used as the baseline (June 2014) and to quantify subsequent yearly relative changes by subdistrict due to migration (Table 3). We speculated that WorldPop estimates might not accurately capture forced displacement, since many Yemeni IDPs live in rented accommodation or communal buildings [14] that would not appear changed in remotely sensed observations.

#### 2.2.2 Internal displacement data

Available displacement datasets covered both IDPs and ‘returnees’ (IDPs who have returned to their communities of origin). Returnee data are acknowledged to feature underestimation [15]. Data were also classifiable as ‘prevalent’ (i.e. information on IDPs or returnees present at a specific time in a given location) or ‘incident’ (novel displacements or returns). Prevalent data sources consisted of baseline or repeat site assessments carried out by the UNHCR-Population Movement Tracking (PMT; 2015-2016) and the International Organisation for Migration (IOM)’s Displacement Tracking Matrix (DTM; 2016-2018) projects. Assessments were sometimes done in-person by agency staff, but largely relied on a network of key informants working with standard templates [16]. The November 2018 assessment round achieved the highest geographical coverage (Supplementary file). During 2018, the IOM also collected incident data on displacements due to insecurity in Al Hudeydah governorate. Since 2019, only incident data have been published.

Displacement datasets were available as unprotected Microsoft Excel worksheets on the IOM DTM site (https://displacement.iom.int/yemen); variable sets, names and formats changed repeatedly over time. We retained the following variables, as available: date of displacement or return; date of assessment (prevalent data only); governorate, district, subdistrict, locality name, geographic codes (hereafter, geocodes) of these administrative levels, and coordinates of the location of arrival / refuge; governorate, district and subdistrict of origin, or of last displacement for returnees, with their geocodes; and number of households. Locality geocodes had an 11-digit structure: digits 1-2 identify the governorate, 1-4 the district and 1-6 the subdistrict. We validated recorded geocodes against the OCHA and CSO gazetteers [6] of all place names (for localities); we also used available locality geocodes to work out missing governorate, district or subdistrict geocodes.

We appended all displacement datasets into one. After applying range and consistency checks, and deleting duplicate records (these were only identifiable for prevalent data, based on identical dates of assessment, displacement/return and locality geocode), the appended dataset consisted of 222,069 records, of which 206,109 (92.8%) concerned IDPs and 15.960 (7.2%) returnees; 195,477 (88.0%) were prevalent-type data. Only IDP data were carried into further analysis, as we assumed returnee data were too incomplete. After removing 2784 (1.3%) records with missing year or month of displacement and 13,188 (6.4%) with missing district of origin or arrival, we retained 190,137 IDP records.

#### 2.2.3 Predictors of displacement

We used multivariate predictive models to impute missing subdistrict data and quantify IDP movements after displacement (see below). While some predictors were built from the population and displacement data themselves, we searched for additional candidate predictor datasets available at month-year and subdistrict resolution. These included (i) a CSO geospatial dataset of Yemen’s road network [17], which we transformed into road density (Km per Km^2^ area); (ii) a crowd-sourced dataset of health facilities [18], which we combined with WorldPop data to estimate health facility density per 100,000 inhabitants; and (iii) the Armed Conflict Location and Event Data Project (ACLED) as a source of georeferenced insecurity event information [19]. Since 2015, ACLED has carried out particularly intensive data collection on Yemen through media monitoring and networks of in-country civil society sources [20]. We mapped each insecurity event to subdistricts based on the event’s coordinates (87/62,629 or 0.1% of records did not map to a subdistrict and were excluded), and aggregated data by subdistrict-month.

### 2.3 Managing the displacement dataset

#### 2.3.1 Standardising place names

Place names in the displacement dataset were a mixture of Arabic and inconsistent Latin character transliterations, and only a fraction of data had unique geocodes, with some geocodes mapping to places that differed from those recorded. We therefore came up with equivalences, down to subdistrict level, between the displacement dataset and the OCHA gazetteer. The dataset featured 5833 unique instances (‘sets’) of missing or non-missing governorate, district, subdistrict names and geocodes. We identified OCHA gazetteer matches for each such set based on the following sequentially applied criteria: (1) the recorded geocode sets (e.g. ‘142024’ for a set down to subdistrict level; ‘1115’ for a set down to district level) also existed in the OCHA gazetteer, and the OCHA place names they mapped to (e.g. governorate 14, Al Bayda, district 1420, Al Malajim and subdistrict 142024, Dhi Khirah) were the same within two characters as the names recorded on the dataset; (2) geocodes were missing, but the place name sets matched a place name set in the OCHA gazetteer within two characters, after applying eight approximate character string matching techniques (stringdist package [21]) and choosing the most common match; (3) the recorded geocode sets also existed in the OCHA gazetteer, and at least the recorded governorate and district names matched with the OCHA names corresponding to the same geocodes (a less stringent version of criterion 1); and (4) a combination of approximate string matching and manual searches applied to any remaining unmatched sets, with matches established at governorate, then district, then subdistrict level so as to restrict each successive search to place names within the same higher-level administrative unit. Criteria 1, 2, 3 and 4 led to a match for 69.2% (4038/5833), 17.3% (1012/5833), 6.5% (380/5833) and 6.9% (403/5833) of unique instances, respectively. All instances were successfully matched.

#### 2.3.2 Imputing missing subdistrict data

After applying the above equivalence, the subdistrict was still missing for 25.6% (48,642/190,137) locations of arrival (31.5%, 30.0%, 1.2%, 49.4%, 0.0% and 100.0% for data collection years 2016 to 2021, respectively) and 79.5% (151,144/190,137) locations of origin (97.8%, 98.7%, 19.1%, 100.0%, 100.0% and 100.0%). Missingness was higher for incident (45.2% and 100.0% for locations of arrival and origin, respectively) than prevalent (22.6% and 76.3%) data. We identified most missing subdistricts through four sequential steps, applied to the individual records (Figure 1):

**Figure 1.**
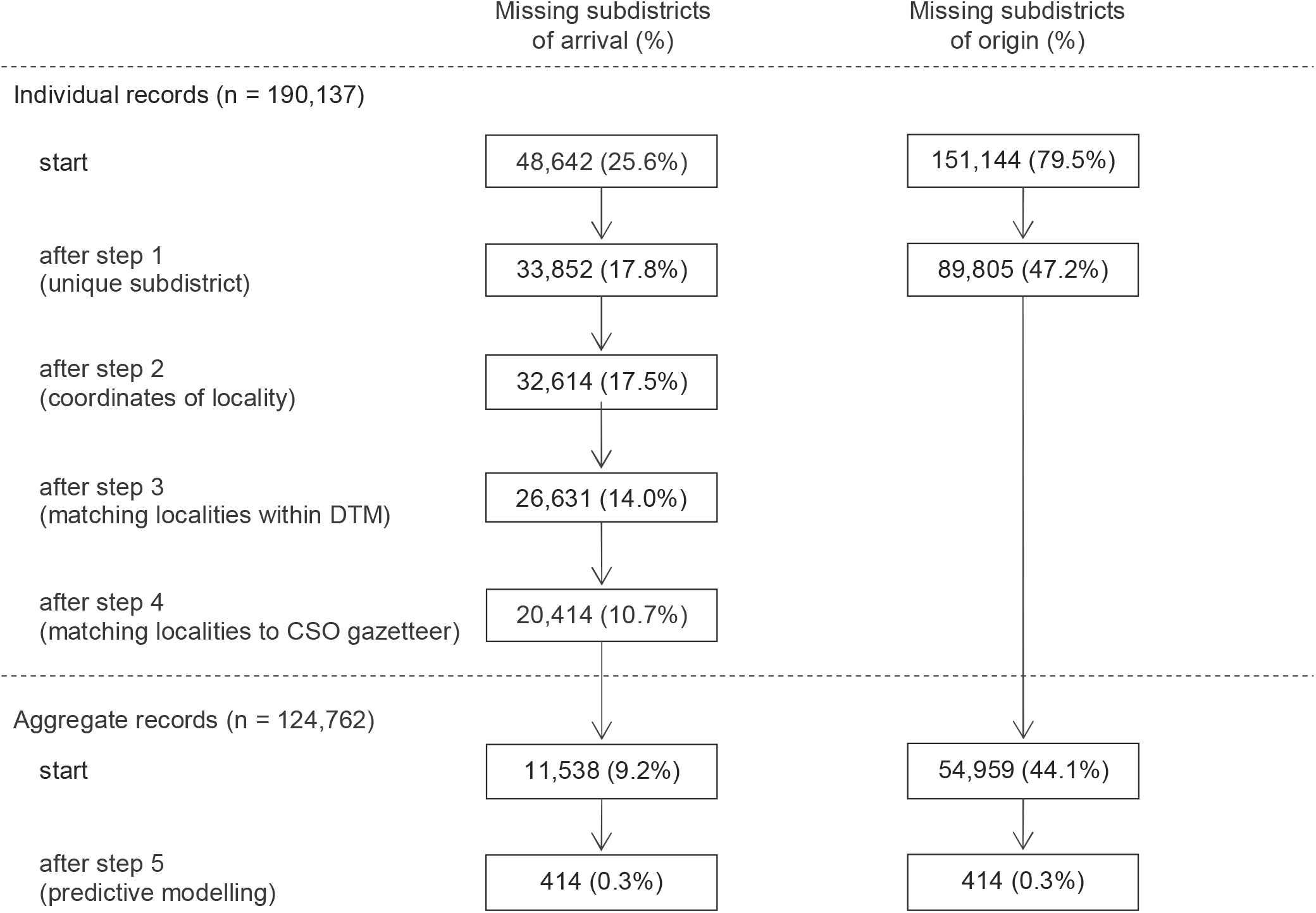
Reductions in subdistrict missingness achieved after each successive data management step. Only eligible IDP records are included in the denominator.

1. Many (n = 117) districts consisted of only one subdistrict;
2. For records that featured longitude and latitude, we identified the subdistrict based on the OCHA/CSO administrative boundaries that the coordinates fell within;
3. We used the above string matching techniques to match the locality name (below subdistrict level), if recorded, to the locality name of other records within the displacement dataset, restricting the matching search to the same governorate and district; if any of the matching localities had a non-missing subdistrict, the latter was applied to matching records with missing subdistrict;
4. We also matched the recorded locality name to locality names at administrative levels below subdistrict (city, neighbourhood, *harrah*, village, sub-village) in the CSO gazetteer, again restricting the matching search to within the same governorate and district, adopting the closest match across the above administrative levels and looking up the subdistrict the CSO match fell within.

We resolved additional missingness through machine learning models. We first predicted how many subdistricts IDPs came from or went to, out of all possible subdistricts in each ‘parent’ district of origin/arrival, by month-year of displacement (Model 1); we then predicted which specific subdistricts IDPs came from or to (Model 2), and the relative share of all IDPs that came from/went to each such subdistrict, out of the parent district total (Model 3). All models were trained on DTM data from November 2018 (n = 41,375), which had 89.8% subdistrict of origin and 100.0% subdistrict of arrival completeness. Training data were aggregated by month-year of displacement, subdistrict of arrival and subdistrict of origin, and augmented to feature all other subdistricts of origin/arrival within the same district, with outcome = 1 attributed to the subdistricts of origin/arrival that any IDPs did come from, and 0 otherwise. For Model 1, training data were further aggregated by district of arrival or origin. For Model 3, training data excluded subdistricts that IDPs did not come from/move to as well as districts with a single subdistrict of origin/arrival.

For each model, random forest algorithms were grown using the ranger R package [22] (weighted for class imbalance and tuned to 500 trees, up to 3 variables to split each node on and maximum tree depth of 20), using the following candidate predictors (at district level for model 1; at subdistrict level otherwise): total population, distance between the geodesic centroids of the subdistricts of origin and arrival, cumulative incidence per capita of insecurity events and fatalities during the current and previous month, health facilities per capita, road density per surface area, surface area, number of candidate subdistricts of origin/arrival within the parent district and the natural log of the number of IDP households from/to the parent district. We evaluated models’ performance out-of-sample using ten-fold cross-validation.

For subdistricts of origin, model 1 yielded fair predictions for the single subdistrict category, but was downward-biased for multi-subdistrict observations (Table 1). As nearly all (97.3%) instances in the training data had only one subdistrict of origin, we applied a simplifying assumption that, for any district of origin - time - subdistrict of arrival combination, all IDPs came from a single subdistrict. For subdistricts of arrival, model 1’s performance was reasonable (Table 1), and we applied the corresponding predictions. Model 2, constrained to the number of subdistricts predicted by model 1, correctly guessed ∼80% of the true subdistricts of origin and ∼63% of true subdistricts of arrival, while incorrectly classifying ∼4% of subdistricts that IDPs did not come from and ∼3% of subdistricts that IDPs did not move to. For model 3, we categorised the outcome (percent share of IDPs) into five classes from 1-19% to 80-99%, with the predicted value at the mid-point of each class. As shown in Table 2, the model had low predictive performance for both origin and arrival subdistricts, though it performed better than a random guess. This low performance was inconsequential for subdistricts of origin (since we assumed no multi-subdistrict instances) and of minor influence for subdistricts of arrival, since model 1 predicted multi-subdistrict instances for only 12.2% of the dataset. Further, we scaled model 3’s predictions to ensure a denominator of 100% for each district.

**Table 1.** Confusion matrices summarising the performance of model 1 to correctly guess the number of subdistricts that IDPs came from or went to, out of all subdistricts within the parent district of origin/arrival. Cell percentages are column-wise. For any observed category, a random guess based only on the category frequencies is shown.

| Model to predict the number of different subdistricts of <u>origin</u> of IDPs within each parent district of origin (not used) |  |  |  |  |  |  |  |  |  |  |  |
| --- | --- | --- | --- | --- | --- | --- | --- | --- | --- | --- | --- |
| Predicted category | Observed category |  |  |  |  | Random guess based on category frequency: |  |  |  |  |  |
|  | 1 | 2 | 3 | 4 | ≥ 5 |  |  |  |  |  |  |
| 1 | 93.9% | 44.7% | 22.5% | 25.0% | 9.5% | 97.3% |  |  |  |  |  |
| 2 | 5.2% | 41.2% | 43.4% | 40.6% | 14.3% | 2.1% |  |  |  |  |  |
| 3 | 0.6% | 10.2% | 22.5% | 12.5% | 33.3% | 0.4% |  |  |  |  |  |
| 4 | 0.2% | 2.8% | 7.0% | 9.4% | 4.8% | 0.1% |  |  |  |  |  |
| ≥ 5 | 0.1% | 1.1% | 4.7% | 12.5% | 38.1% | 0.1% |  |  |  |  |  |
| Model to predict the number of different subdistricts of <u>arrival</u> of IDPs within each parent district of arrival |  |  |  |  |  |  |  |  |  |  |  |
| Predicted category | Observed category |  |  |  |  |  |  |  |  |  | Random guess based on category frequency: |
|  | 1 | 2 | 3 | 4 | 5 | 6 | 7 | 8 | 9 | ≥ 10 |  |
| 1 | 81.4% | 19.6% | 2.7% | 0.6% | 0.6% | 0.0% | 0.0% | 0.0% | 0.0% | 0.0% | 82.7% |
| 2 | 14.9% | 52.8% | 32.9% | 15.9% | 5.3% | 2.1% | 1.5% | 0.0% | 0.0% | 0.0% | 10.4% |
| 3 | 2.8% | 17.7% | 36.7% | 30.3% | 14.2% | 7.3% | 5.9% | 0.0% | 0.0% | 0.0% | 3.6% |
| 4 | 0.6% | 6.5% | 16.5% | 27.4% | 22.5% | 15.6% | 1.5% | 13.6% | 9.1% | 0.0% | 1.5% |
| 5 | 0.2% | 1.9% | 6.7% | 12.7% | 27.2% | 21.9% | 8.8% | 9.1% | 13.6% | 1.7% | 0.7% |
| 6 | 0.1% | 0.6% | 1.4% | 6.9% | 13.6% | 14.6% | 22.1% | 6.8% | 22.7% | 3.3% | 0.4% |
| 7 | 0.0% | 0.1% | 0.7% | 0.9% | 4.7% | 14.6% | 26.5% | 22.7% | 13.6% | 5.0% | 0.3% |
| 8 | 0.0% | 0.4% | 0.5% | 2.9% | 4.7% | 8.3% | 14.7% | 18.2% | 22.7% | 11.7% | 0.2% |
| 9 | 0.0% | 0.2% | 1.4% | 2.0% | 5.3% | 9.4% | 8.8% | 13.6% | 0.0% | 13.3% | 0.1% |
| ≥ 10 | 0.0% | 0.0% | 0.5% | 0.6% | 1.8% | 6.2% | 10.3% | 15.9% | 18.2% | 65.0% | 0.3% |

**Table 2.** Confusion matrices summarising the performance of a random forest model to correctly guess the percent share of IDPs coming from or to a given subdistrict (expressed as a categorical variable), out of all IDPs from or to the parent district. Cell percentages are column-wise. For any observed category, a random guess based only on the category frequencies is shown.

| Model to impute percent share of IDPs by subdistrict of <u>origin</u> (not used) |  |  |  |  |  |  |
| --- | --- | --- | --- | --- | --- | --- |
| Predicted category | Observed category |  |  |  |  | Random guess based on category frequency: |
|  | 1 to 19% | 20 to 39% | 40 to 59% | 60 to 79% | 80 to 99% |  |
| 1 to 19% | 43.4% | 27.5% | 22.5% | 21.9% | 23.1% | 21.6% |
| 20 to 39% | 21.3% | 22.9% | 19.2% | 23.8% | 16.0% | 27.5% |
| 40 to 59% | 12.0% | 17.7% | 28.5% | 21.6% | 14.1% | 25.4% |
| 60 to 79% | 10.0% | 20.2% | 19.0% | 16.7% | 19.2% | 17.2% |
| 80 to 99% | 13.2% | 11.7% | 10.8% | 16.0% | 27.6% | 8.3% |
| Model to impute percent share of IDPs by subdistrict of <u>arrival</u> |  |  |  |  |  |  |
| Predicted category | Observed category |  |  |  |  | Random guess based on category frequency: |
|  | 1 to 19% | 20 to 39% | 40 to 59% | 60 to 79% | 80 to 99% |  |
| 1 to 19% | 61.30% | 22.70% | 15.30% | 12.30% | 21.00% | 60.2% |
| 20 to 39% | 16.30% | 32.10% | 25.70% | 32.90% | 21.40% | 24.3% |
| 40 to 59% | 8.30% | 15.30% | 31.60% | 8.90% | 15.20% | 10.6% |
| 60 to 79% | 9.90% | 25.50% | 22.10% | 38.90% | 30.80% | 4.1% |
| 80 to 99% | 4.10% | 4.30% | 5.30% | 6.90% | 11.60% | 0.8% |

Overall, we imputed missing subdistricts for all but a small minority of displacement records (Figure 1), which were excluded from further analysis.

### 2.4 Population reconstruction

#### 2.4.1 General equations

Let *N* be a two-dimensional matrix with dimensions *i* ∈ (1, 2, 3 … / substricts of arrival) and *t* ∈ (1, 2, 3 … *T*, with 1 = June 2014 and *T* = September 2021, and each increment = one month) where *n*_*i t*_is the population of subdistrict *i* at the start of month t. Let F be a three-dimensional matrix of forced displacement, with dimensions *t, i* and *j* ∈ (1, 2, 3 … ] subdistricts of origin), where *f*_*i jt*_is the net flow of IDPs from j to i during month t (IDPs can be displaced within their subdistrict, in which case *i* = *j)*. We further define B, D and M as matrices with dimensions *i* and *t* where *b*_*it*_and *d*_*it*_are birth and death rates per capita during each subdistrict-month, *and m*_*it*_is the percent change in subdistrict *i*’s population during month *t* resulting from migration *other than* forced displacement. Lastly, *φ* is the proportion of forced displacement that is already taken into account by M, i.e. the extent to which data on migration also capture forced displacement (if *φ*= 0, the two data sources have no overlap; cp = 1 implies complete overlap). It follows that

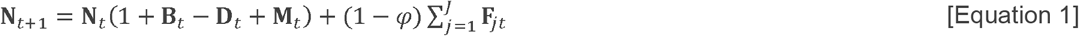

Otherwise put, the following month’s population is this month’s starting population multiplied by the net rate of natural growth and migration, plus any net change in IDPs that isn’t already captured by migration estimates.

#### 2.4.2 Estimating displacement flows

To populate matrix F, we needed to combine prevalent and incident data. We aggregated all data to identify unique IDP groups that moved from a given subdistrict of origin to a given subdistrict of refuge during a given month (we call these ‘instances’, denoting discrete waves of primary displacement). Each such instance was subject to one or more longitudinal observations (DTM site assessments). For a minority (30.2% or 6258/20,744) of multi-observation instances, later observations in the dataset featured a higher number of IDPs than at previous assessment points, which is theoretically impossible beyond marginal increases due to natural growth. Most such cases were moderate and occurred when small numbers of IDP households were involved. We assumed these were due to clerical error, data collection problems or the discovery of previously undetected IDP households. We converted these problematic instances into constant or monotonically decreasing series based on alternative assumptions (Table 3).

**Table 3.** Input values for parameters, by analysis.

| Parameter (symbol) | Main analysis | Reasonable-low sensitivity scenario | Reasonable-high sensitivity scenario |
| --- | --- | --- | --- |
| Monthly flow of IDP households from/to subdistricts (F) | Prevalent-data instances in which the reported number of IDP households increased from the previous assessment point were adjusted as follows: |  |  |
|  | Average of reasonable-low and reasonable-high adjusted values (see columns to the right). | All values higher than the previous value were changed to the previous value. | All values lower than the next value were changed to the next value. |
|  | Monthly prevalence predictions were based on the additive growth model (see text). The predicted series was then converted to incident flows by computing month-on-month changes |  |  |
| Mean number of IDPs per household | 6.70 (countrywide estimate) [24] | 5.34 (assume -20%) | 8.04 (assume +20%) |
| Population of each subdistrict at the base time point, June 2014 ($N_{t=\text{Jun } 2014}$) | WorldPop estimates, corrected for prevalent displacement at that time point. | As for main analysis, but using $F_{\text{low}}$ | As for main analysis, but using $F_{\text{high}}$ |
| Relative change per month due to migration (M) | First, we adjusted annual WorldPop estimates for 2014-2020 by eliminating UN-projected natural growth (2.8% per annum). Then, we inter- and extrapolated annual estimates using a natural cubic spline to obtain monthly values $u_{it}$. Lastly, computed $m_{it} = \frac{u_{it} - u_{i(t-1)}}{u_{i(t-1)}}$, starting with $t = \text{Jun } 2014$. | As for main analysis | As for main analysis |
| Proportion of overlap between data on migration changes and displacement data ($\varphi$) | 0.25 | 0.50 | 0 (no overlap) |
| Crude birth rate (B) | First, performed smooth interpolation of secular trends according to UN World Population Prospects to obtain an assumed monthly value in the absence of a crisis. Second, came up with value for rural and urban subdistricts based on the ratio of crude birth rate observed in the last DHS survey [24], weighted for relative population as of June 2014. Third, applied a multiplier to model crisis- and COVID-19 attributable changes (see Figure 4). For main analysis, assumed extension of pre-crisis trend. | See Figure 4. Assumed that the crisis would result in a sudden drop in fertility. | See Figure 4. Assumed that the crisis would disrupt family planning. |
| Crude death rate (D) | Same approach as for birth rate (see Figure 4). Ratio of urban to rural crude death rate based on the ratio of under 5y mortality in the last DHS survey [24]. For main analysis, assumed progressive increases based on timeline of crisis intensity and onset of COVID-19 pandemic [25]. Indicatively, countrywide deaths were estimated to increase by 1.15 during Somalia's 2016-2018 drought [26] and by 1.42 following the 2003 Coalition invasion in Iraq [27]. | See Figure 4. Assumed. | See Figure 4. Assumed. |

As depicted in Figure 2, prevalent observations may underestimate past displacement, since during the period before assessments all or some of the IDPs may have returned home or moved to another subdistrict. This bias is dependent on the rate of return or onward movement. Equally, how IDP populations evolve after the last assessment is unknown.

**Figure 2.**
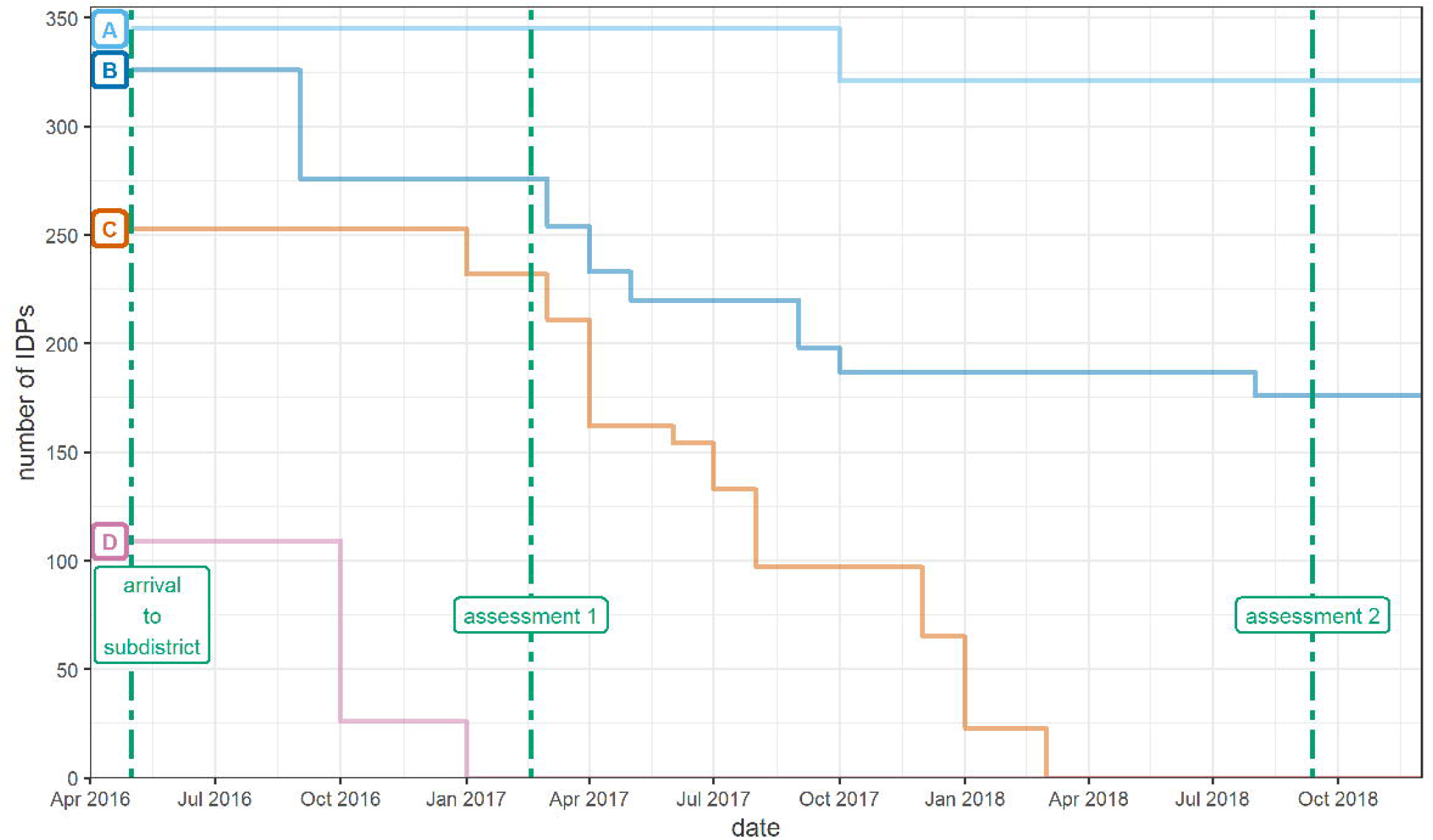
Illustration of four hypothetical scenarios (A to D) in which a group of IDPs arrives to a subdistrict from another subdistrict. In scenario A, nearly all IDPs remain in the subdistrict of refuge throughout the period of interest. In scenario B, only a fraction are left by the second assessment round. In scenario C, all IDPs have left the subdistrict (either returned to their subdistrict of origin, or moved elsewhere) by the second assessment, and in scenario D IDPs have left even before the first assessment, thereby potentially being missed altogether by the displacement tracking system.

To quantify this evolution and thereby predict IDP populations before and after the timeframe of available observations, we used the gamlss framework [23] to fit a generalised additive mixed growth model to all unique instances, as defined above, for which at least two prevalent assessment observations existed, excluding records for which the subdistrict was imputed. The model predicted the number of IDP households as a monotonic penalised-spline smoothed function of time since displacement, with IDP instance as a random effect, and, as predictors, distance between subdistricts of origin and arrival, health facility density (arrival), road coverage (origin and arrival) and a monotonic spline of insecurity event incidence in the subdistrict of origin since the previous assessment. We assumed a quasi-Poisson distribution for the data, as this provided reasonable model diagnostics. Figure 3 suggests that, on average, about 75% of IDPs left the subdistrict of arrival within two years of first displacement; thereafter, departures reduced.

**Figure 3.**
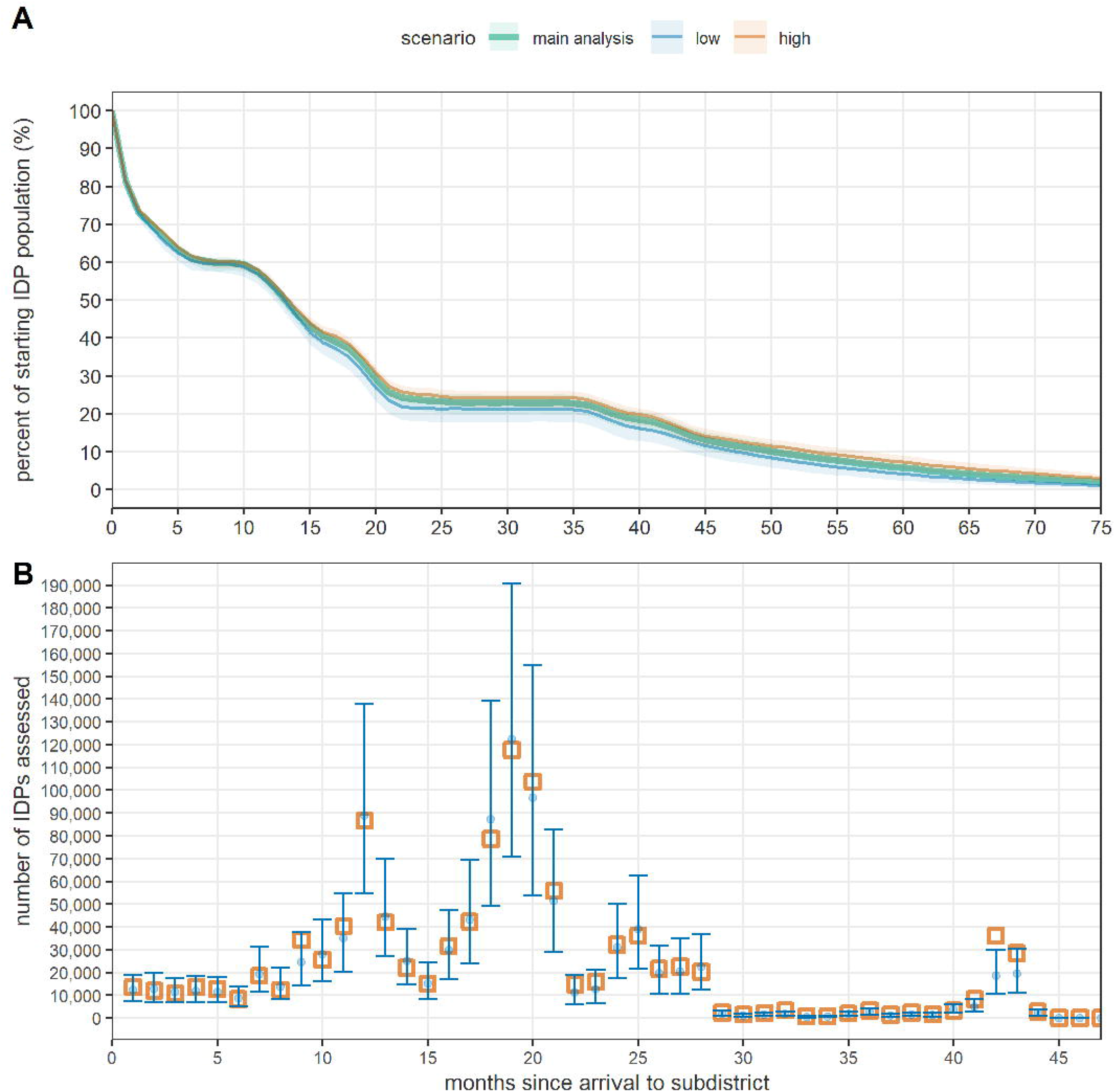
Predictions of a generalised additive mixed growth model of IDP population as a function of time. Panel A shows the model’s predicted percent change in IDP group size by month since displacement: the thick line is the main analysis prediction, and the shaded area indicates the 95%CI. Predictions using reasonable-high and -low adjustments for non-monotonic series are also shown. Panel B shows the model’s predictions (blue dots and 95% confidence bands) and observed total prevalent number of IDPs assessed (orange squares) during any given month after displacement, for the main analysis only.

We used the model to predict the evolution of IDP household counts across time for both the model-training data instances and all other (i.e. single-assessment and incident) instances: for the latter, we made predictions using only the fixed-effects model coefficients, and scaled these to the single recorded IDP household count. We made a simplifying assumption that IDPs either stayed in the subdistrict of first arrival or returned to their subdistrict of origin (i.e. zero secondary displacement).

#### 2.4.3 Input values

Input values for all equation parameters are detailed in Table 3 and Figure 4.

**Figure 4.**
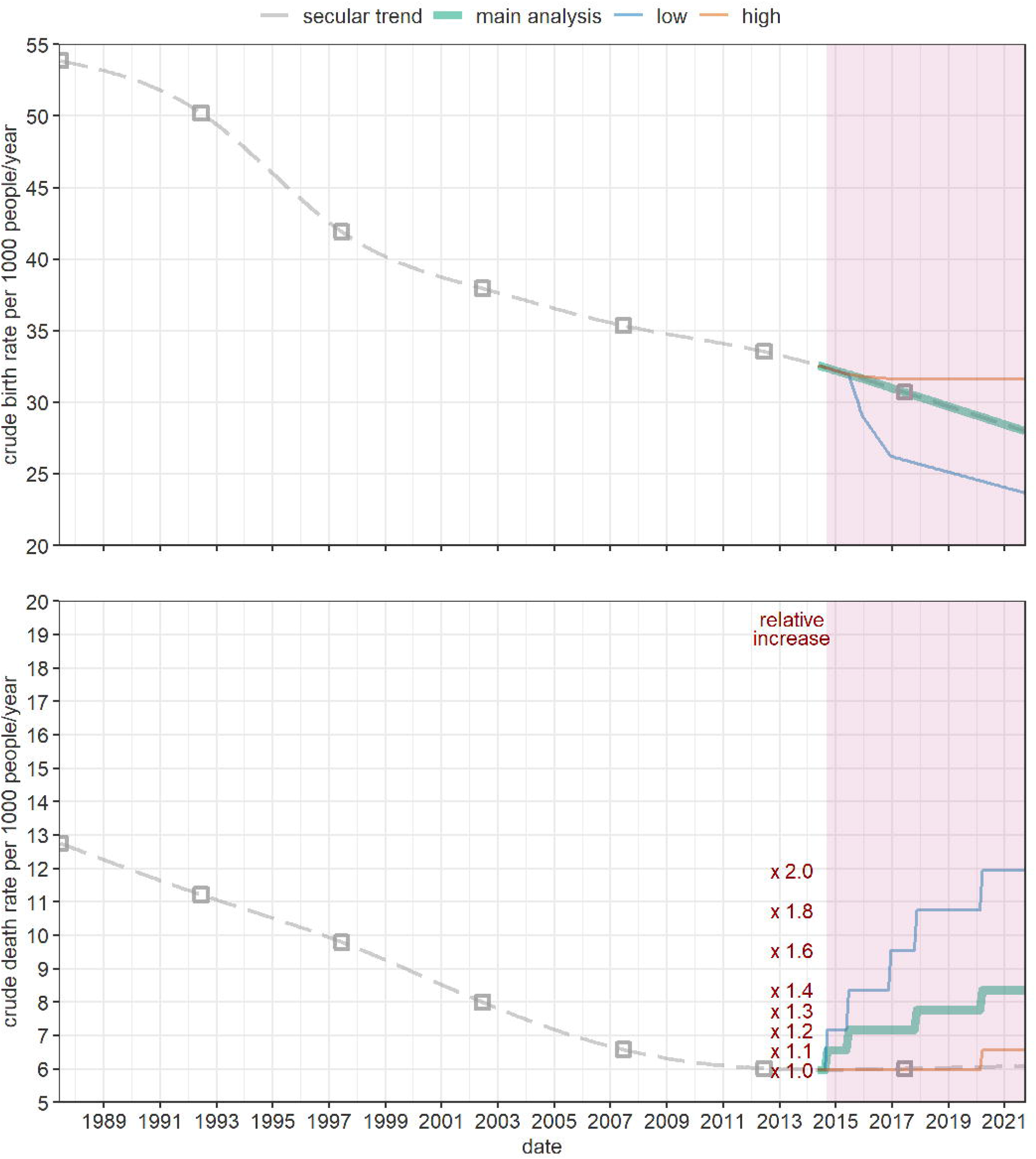
Countrywide average values of crude birth rate and crude death rate assumed, by scenario. The grey dotted line shows secular trends based on UN projections; the latter are represented by squares and centred at the mid-point of their period of reference. The shaded area indicates the crisis period.

**Table 4.** Estimated population by governorate as of September 2021, and percent change from the June 2014 baseline. Figures include the main analysis and, in parentheses, and reasonable-low and -high scenarios.

| Governorate | Estimated population (Sep 2021) | Percent change from Jun 2014 |
| --- | --- | --- |
| Abyan | 703,000 (665,000 to 729,000) | 22.4 (15.6 to 26.9) |
| Ad Dali' | 759,000 (721,000 to 781,000) | 20.8 (14.7 to 24.1) |
| Aden | 943,000 (911,000 to 956,000) | 17.8 (13.8 to 19.5) |
| Al Bayda | 907,000 (866,000 to 926,000) | 19.1 (13.7 to 21.5) |
| Al Hodeidah | 3,270,000 (3,208,000 to 3,221,000) | 13.7 (11.6 to 12.0) |
| Al Jawf | 680,000 (653,000 to 688,000) | 17.5 (12.8 to 18.7) |
| Al Maharah | 148,000 (139,000 to 156,000) | 24.9 (17.0 to 30.7) |
| Al Mahwit | 807,000 (763,000 to 841,000) | 19.9 (13.4 to 25.0) |
| Amran | 1,501,000 (1,408,000 to 1,595,000) | 20.5 (13.1 to 27.9) |
| Dhamar | 2,188,000 (2,050,000 to 2,304,000) | 22.1 (14.5 to 28.6) |
| Hadramawt | 1,568,000 (1,493,000 to 1,607,000) | 21.0 (15.3 to 24.1) |
| Hajjah | 2,273,000 (2,203,000 to 2,284,000) | 14.7 (11.2 to 15.3) |
| Ibb | 3,412,000 (3,253,000 to 3,534,000) | 17.0 (11.5 to 21.1) |
| Lahj | 1,148,000 (1,085,000 to 1,190,000) | 21.9 (15.2 to 26.4) |
| Ma'rib | 668,000 (483,000 to 925,000) | 108.3 (50.7 to 188.2) |
| Raymah | 629,000 (603,000 to 643,000) | 17.4 (12.4 to 19.9) |
| Sa'dah | 964,000 (996,000 to 846,000) | 5.3 ( 8.8 to -7.5) |
| Sana'a | 1,331,000 (1,279,000 to 1,347,000) | 20.4 (15.8 to 21.9) |
| Sana'a City | 2,723,000 (2,648,000 to 2,729,000) | 15.2 (12.1 to 15.5) |
| Shabwah | 762,000 (725,000 to 782,000) | 20.2 (14.4 to 23.2) |
| Socotra | 70,000 (67,000 to 72,000) | 21.5 (15.5 to 24.5) |
| Ta'iz | 3,699,000 (3,615,000 to 3,666,000) | 13.5 (10.9 to 12.5) |
| **Total** | **31,154,000 (29,835,000 to 31,821,000)** | **18.1 (13.1 to 20.6)** |

## 3 Results

We estimated that Yemen’s population rose from 26,376,000 in June 2014 (of whom 3,571,000 children aged under 5y) to 31,154,000 (4,232,000) in September 2021 (Figure 5). At governorate level, population trends were variable (Figure S11), with very sudden increases or decreases in 2015, coinciding with large-scale displacement (Figure S12).

**Figure 5.**
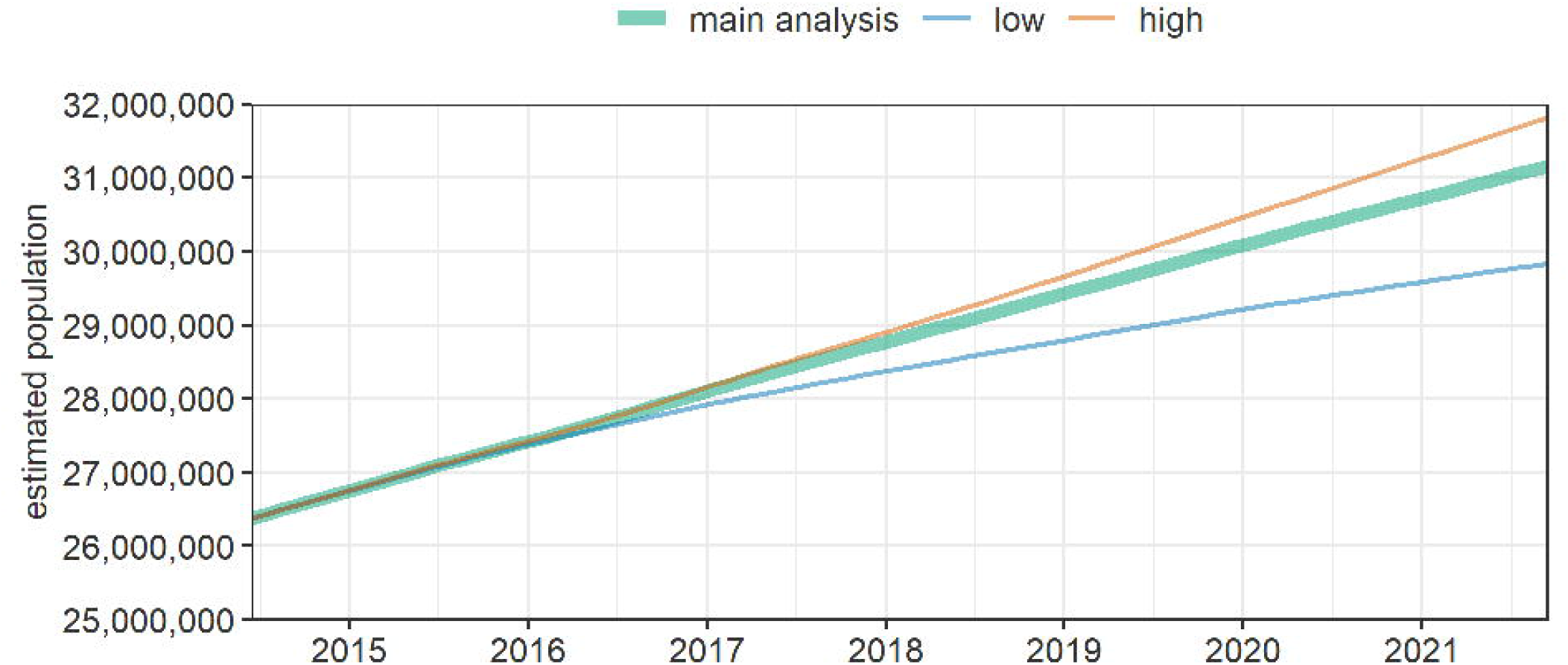
Estimated population of Yemen over time, by scenario.

Relative change from baseline was within the 15-25% range, but we estimated that Ma’rib governorate approximately doubled in size, while Sa’dah experienced minimal growth, reflecting displacement to and from these areas. A negative population estimate occurred for 2.6%, 1.5% and 3.7% of subdistrict-months in the main analysis, reasonable-low and reasonable-high scenarios, respectively.

In contrast to estimates produced by the IOM and the Internal Displacement Monitoring Centre, we estimated that some 10 to 14M Yemenis were displaced during the period from mid-2015 to mid-2016, amounting to some 40% of the population (Figure 6; see Discussion). Our period-end estimate of total IDPs, however, was considerably lower than that reported by the United Nations (between 0.9 and 2.1M versus 3.3M).

**Figure 6.**
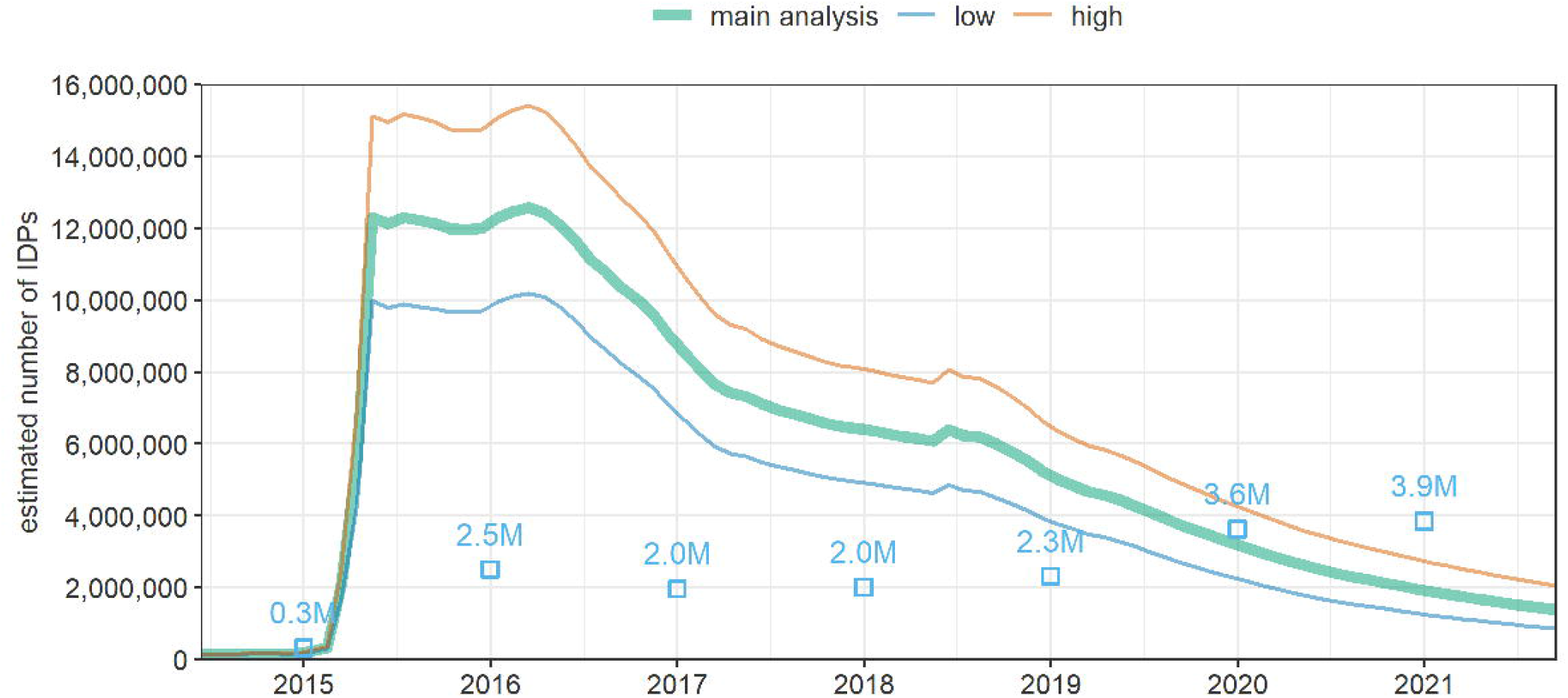
Estimated number of IDPs in Yemen over time, by scenario. Squares with labels (in millions or M) indicate year-end estimates from the Internal Displacement Monitoring Centre [15].

In the majority of districts IDPs were <10% of the overall population (Figure 7), but Ma’rib, Sa’dah and Ta’iz had higher relative presence of IDPs (one district had a negative estimated population and thus no calculable IDP percentage).

**Figure 7.**
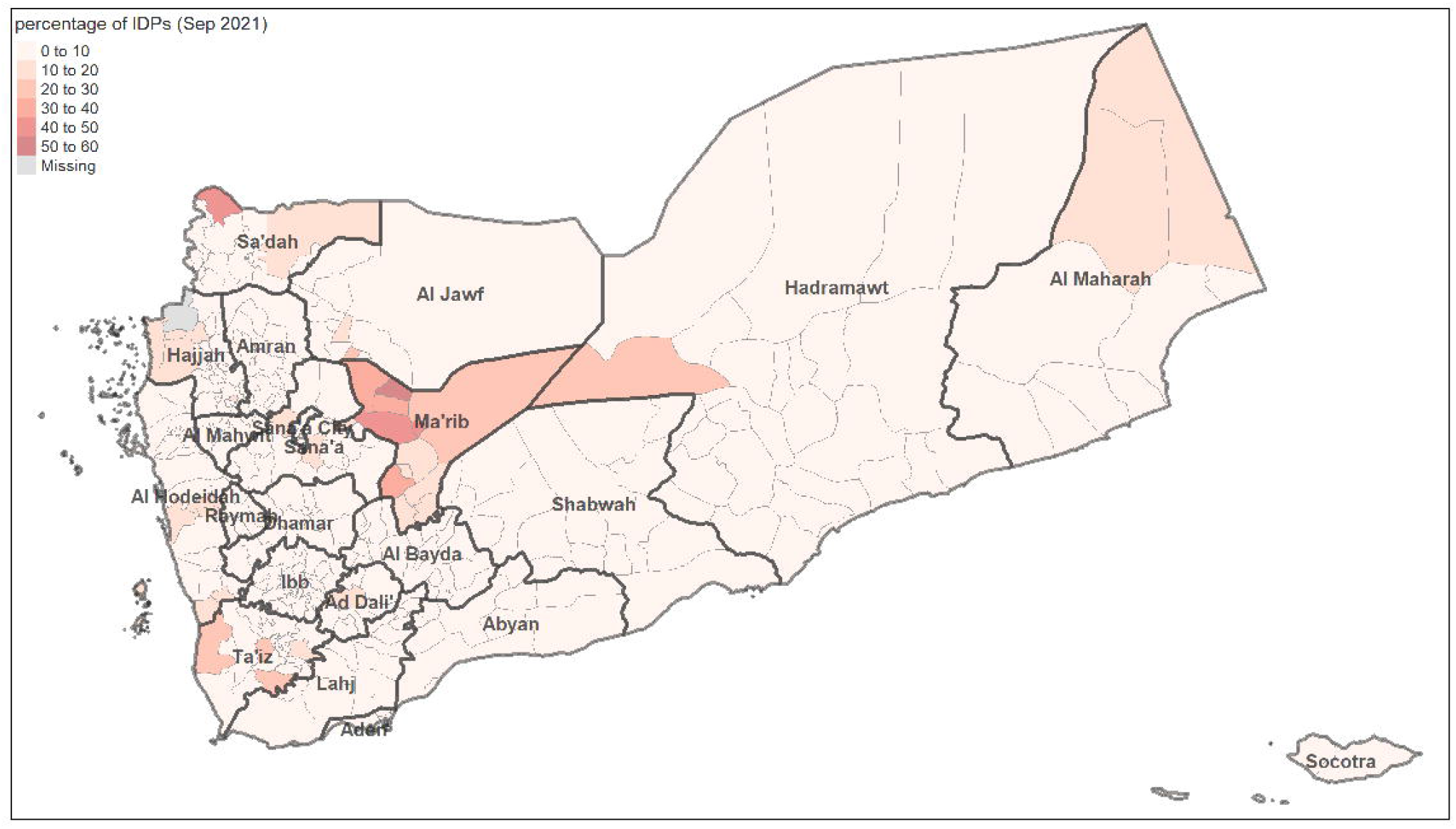
Estimated percentage of IDPs among the entire population, by district, as of September 2021. Thick boundaries and text labels denote governorates; light boundaries denote districts.

## 4 Discussion

To our knowledge ours is the first attempt to reconstruct the demographic evolution of Yemen’s population while taking into account changes due to the past seven years of war and food insecurity. Our analysis finds that the population of Yemen increased by about 3M to 6M over a seven-year period, though displacement and internal migration caused substantial demographic shifts at governorate and lower administrative levels. We estimate that a surprisingly large percentage of Yemen’s population may have been displaced in the early phase of the crisis. Displacement has wide-ranging effects on livelihoods, security, health and child development [28]: its occurrence at such scale suggests that a large number of Yemenis have been deeply affected by the crisis. Models, however, indicate that some primary displacement was short-lived, with some 40% moving on or returning within the first year.

Aside from the estimates themselves, our analysis demonstrates the applicability of various data science methods, including machine learning, to make sense of large but incomplete primary data. In particular, we were able to predict the origin and arrival of IDPs with reasonable accuracy by associating to the DTM records other openly available datasets, including, critically, insecurity. Huynh and Basu [29] have applied similar models for Syria and Yemen to accurately predict future displacement. Data science methods have also been used to predict refugee [30] and migrant [31] destinations, and the timing of migratory flows into Europe [32].

### 4.1 Limitations

Our analysis excludes refugees and other migrants leaving or entering Yemen, though these are expected to be few. It also does not capture secondary displacement, and instead simplistically assumes that IDPs could only move back to their subdistrict of origin; we did not identify any data to realistically explore the sensitivity of findings to this assumption.

We attempted to represent uncertainty in the estimates by featuring reasonable worse- and best-case scenarios. However, many parameters (e.g. birth and death rate, household size) would likely vary considerably at subdistrict level, and furthermore the range of scenarios does not account for error in the different statistical models underlying input data, including the WorldPop geospatial predictions of population density and our own models to impute missing subdistricts and number of IDP households within these. Summation of inaccuracy in the latter models would probably have caused erroneous imputations of subdistricts of arrival and origin for ∼ 4% and ∼ 9% of DTM records, respectively. As model predictions were constrained to fall within the known parent district data, our estimates are not affected by imputation uncertainty at the district or governorate level.

Critically, our analysis is very sensitive to inaccuracy in census data (the basis for countrywide projections that WorldPop estimates are scaled to) and displacement figures: the latter mostly rely on key informant reports rather than ground estimation, and may also entail differential bias by period or location in terms of the proportion of IDP movements captured. We did not have any means of validating these source data, though we noted evidence of rounding and digit preference in reported IDP numbers, suggesting approximations. Our estimates suggest a far greater number of Yemenis were displaced in the early phase of the crisis than official figures suggest: this difference, which may appear implausible, is largely driven by our modelled evolution of IDP populations after displacement. While this model may be inaccurate, it does reflect the general pattern in multi-assessment DTM instances, namely that IDP numbers in any given location declined considerably over time since displacement. Official estimates, by contrast, appear static, with updates only when new prevalent or incident data are available, and/or based on probable underestimates of returnees. The DTM project carried out countrywide assessments with similar district coverage in November 2016 and November 2018 (Figure S9). The former recorded some 2.0M IDPs, far less than our estimate for the same time point, and the latter 3.7M. When considering instances with a displacement date predating both assessments (N = 65,101), we found that 47.6% appeared in the 2016 assessment, 35.4% in the 2018 assessment and only 4.9% in both. Crude two-list capture-recapture analysis based on this contingency table suggests only ∼18% of instances were ever recognised, which is intriguingly close to the ratio of Internal Displacement Monitoring Centre to our estimates (∼21%) around what we project to be the peak period of prevalent displacement (2015-2016). Gallup polls and World Food Programme random phone surveys have suggested that up to a third of Yemenis may have ever been displaced during the crisis [33], roughly consistent with our estimates.

## 5 Conclusions

Ultimately, uncertainty in these and official population and displacement estimates underscores the importance of consistent, well-resourced data collection in crisis settings. The political and security challenges of Yemen’s information landscape have been described [34]. In this context, the IOM and partners’ efforts to collect displacement data with large geographical coverage are laudable. However, a few key adaptations to the DTM could enable far easier and more robust analysis of IDP trends, obviating the need for models. First, DTM data should collect the same set of variables consistently: these should include the location of both origin and arrival (based on the official gazetteer). Groups of IDPs (e.g. camps or clusters of households from the same location) should be attributed a unique identifier, allowing for their tracking over time and enabling estimation (e.g. through capture-recapture methods) of the sensitivity of data collection of each DTM assessment, i.e. of the likely true number of IDPs out there. IDPs themselves could be asked about returns or onwards movements from within the group that they originally travelled with. Such personal data, however, should be collected and managed without incurring security concerns and risks for IDPs themselves. Generally, displacement analysis must be dynamic, i.e. monitor flows and update prevalent estimates accordingly. These improvements may require higher investment by humanitarian donors into the DTM or other systems, but not without concurrent improvements in design and analysis.

Humanitarian response and service planning are unlikely to be appropriate if population denominators are unclear. Inefficiency at best, and avertable mortality at worst, are the likely consequences. Crisis-affected populations must be counted properly as a key starting point for properly supporting them.

## Data Availability

All data produced are available online at https://github.com/francescochecchi/yem_pop_reconstruction .

## 6 Declarations

### 6.1 Ethics approval and consent to participate

Not applicable: all data were in the public domain and contained no unique identifiers.

### 6.2 Consent for publication

Not applicable.

### 6.3 Availability of data and materials

All R analysis scripts and input data are available at https://github.com/francescochecchi/yem_pop_reconstruction.

### 6.6 Authors’ contributions

FC designed the study, collected data, did analysis and wrote the paper. EKB designed the study, collected data and did analysis.

## 6.7 Acknowledgments

We are grateful to the International Organisation for Migration for sharing information on how DTM data were collected, to Mervat Alhaffar for reviewing the manuscript, to Lucy Bell for project management support, to Claire Dooley for advice on WorldPop data and to the Foreign, Commonwealth and Development Office for facilitating contacts with humanitarian actors in Yemen. The Satellite Applications Catapult kindly supplied WorldPop datasets disaggregated by administrative boundary.

