## Supplementary file for "Reconstructing subdistrict-level population denominators in Yemen after six years of armed conflict and forced displacement"

SUPPLEMENTARY MATERIAL

Francesco Checchi PhD*, Emilie Koum Besson

Department of Infectious Disease Epidemiology

Faculty of Epidemiology and Population Health

London School of Hygiene and Tropical Medicine

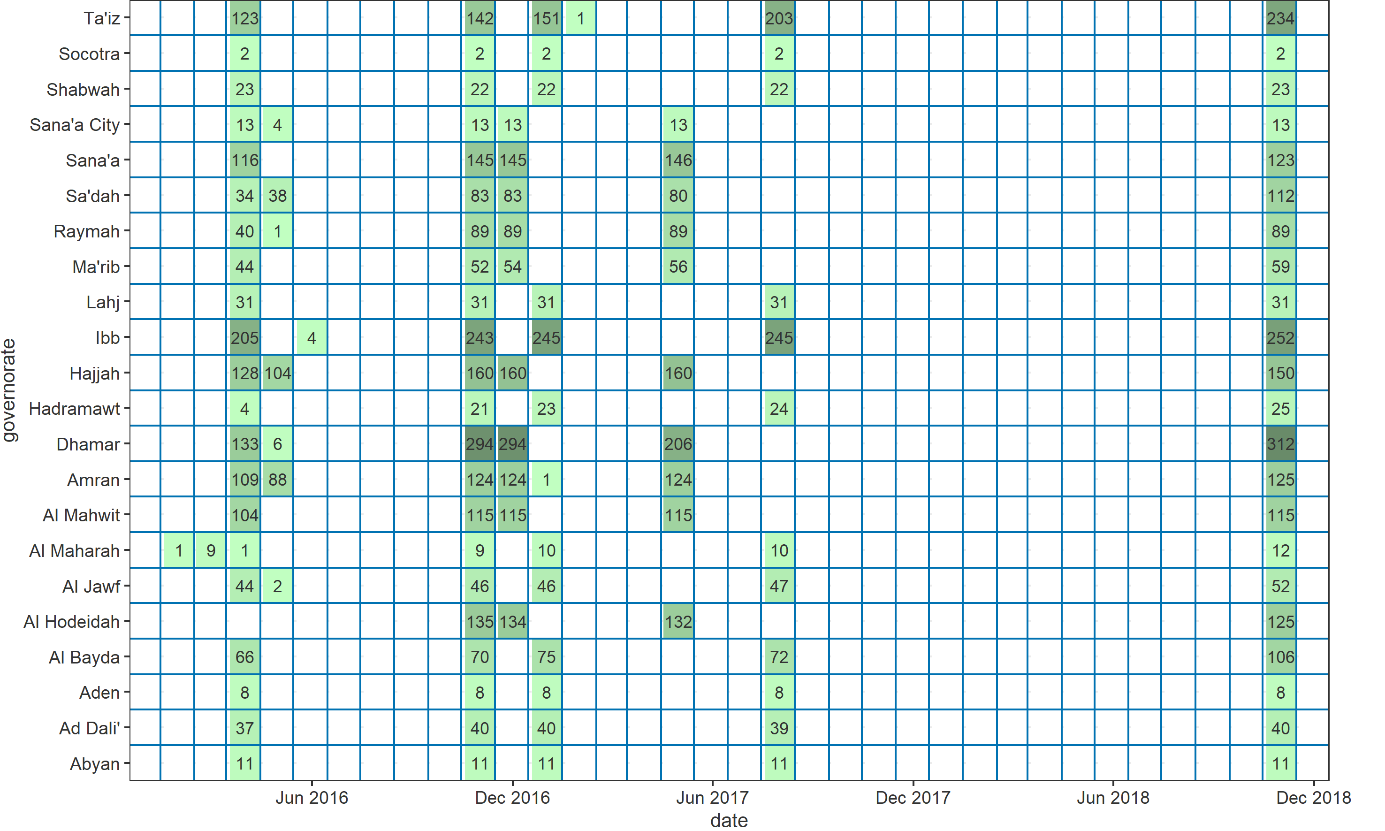

Figure S8. Number of subdistricts of arrival included in prevalent displacement data, by governorate and date of assessment. Includes imputed subdistricts.

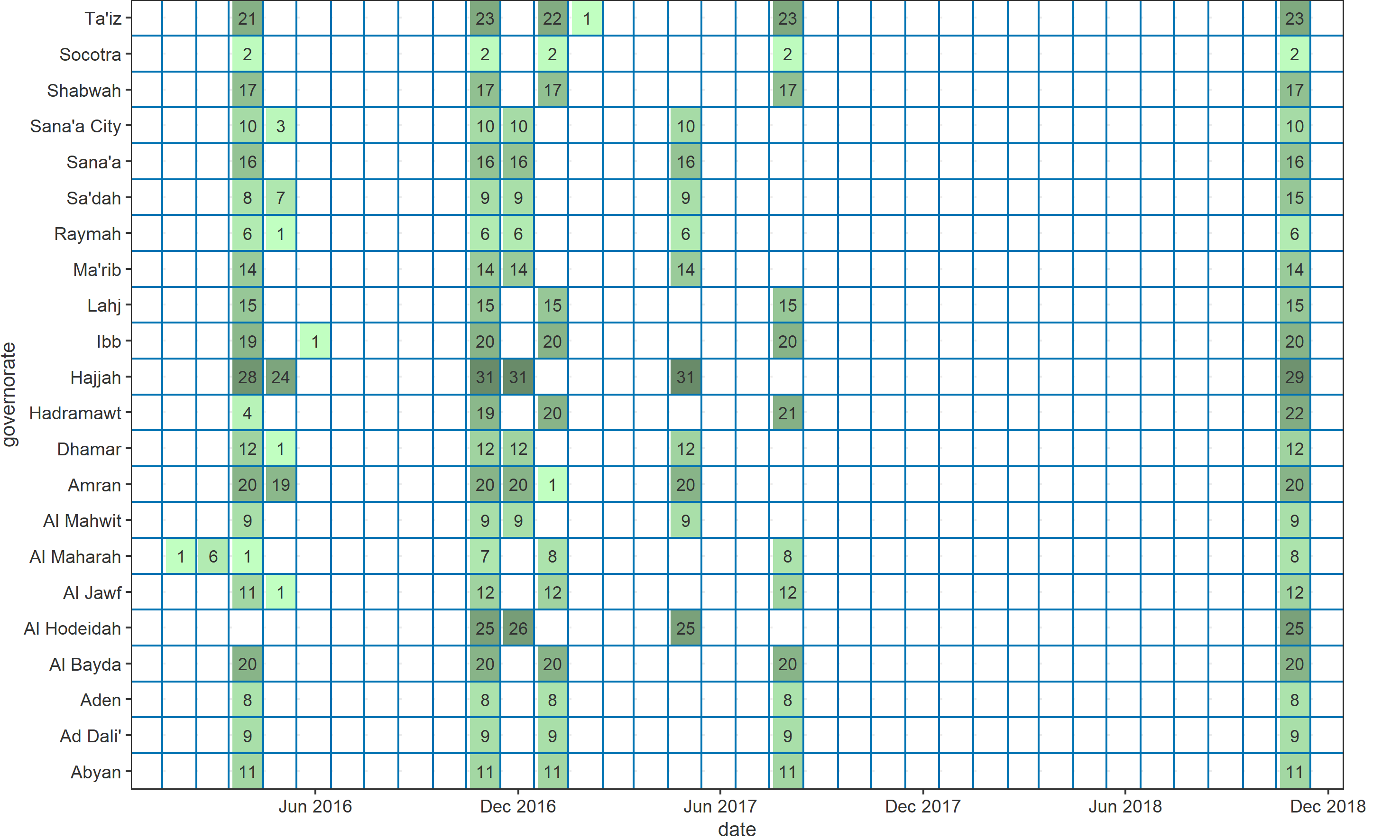

Figure S9. Number of districts of arrival included in prevalent displacement data, by governorate and date of assessment.

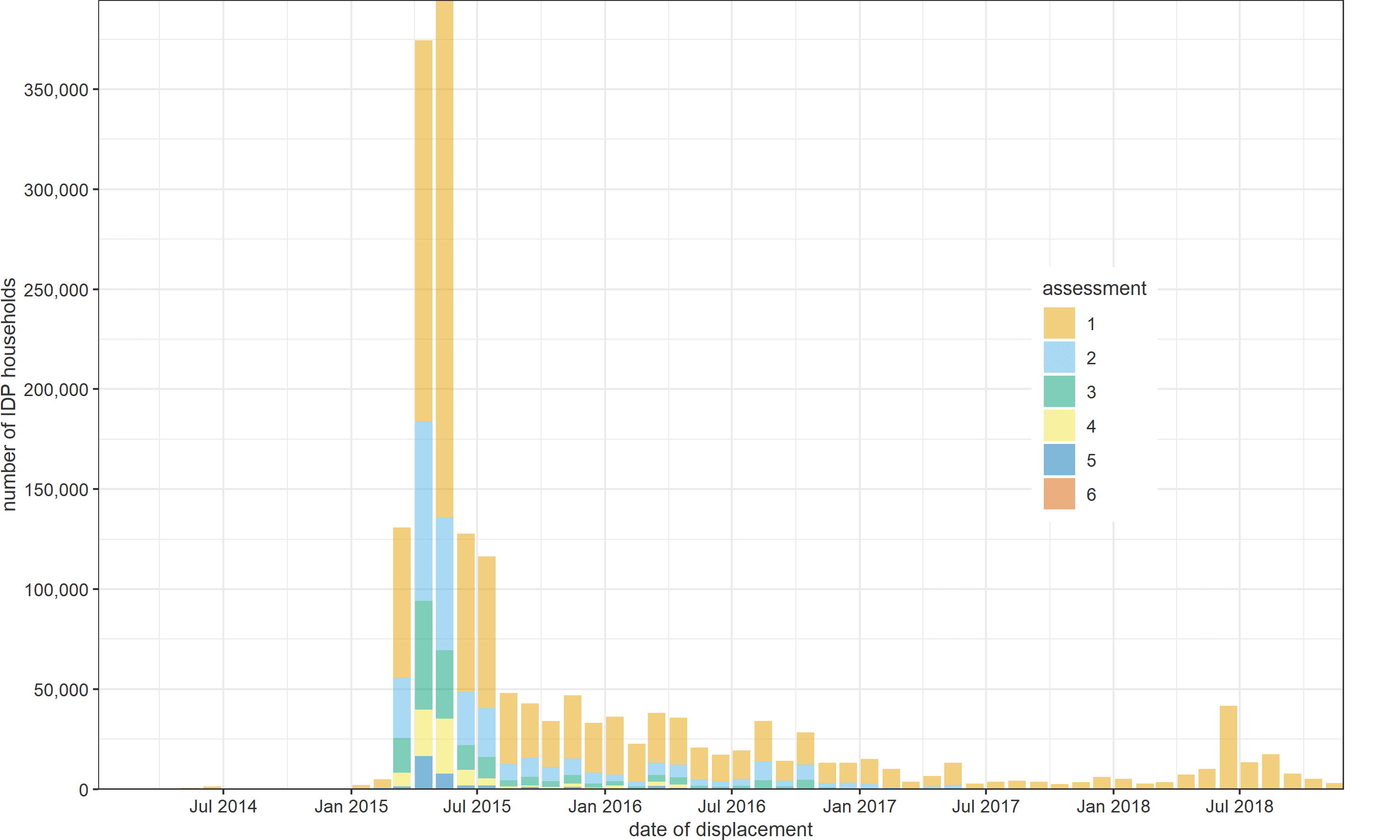

Figure S10. Number of reported IDP households, by recorded month of displacement and DTM prevalent assessment.

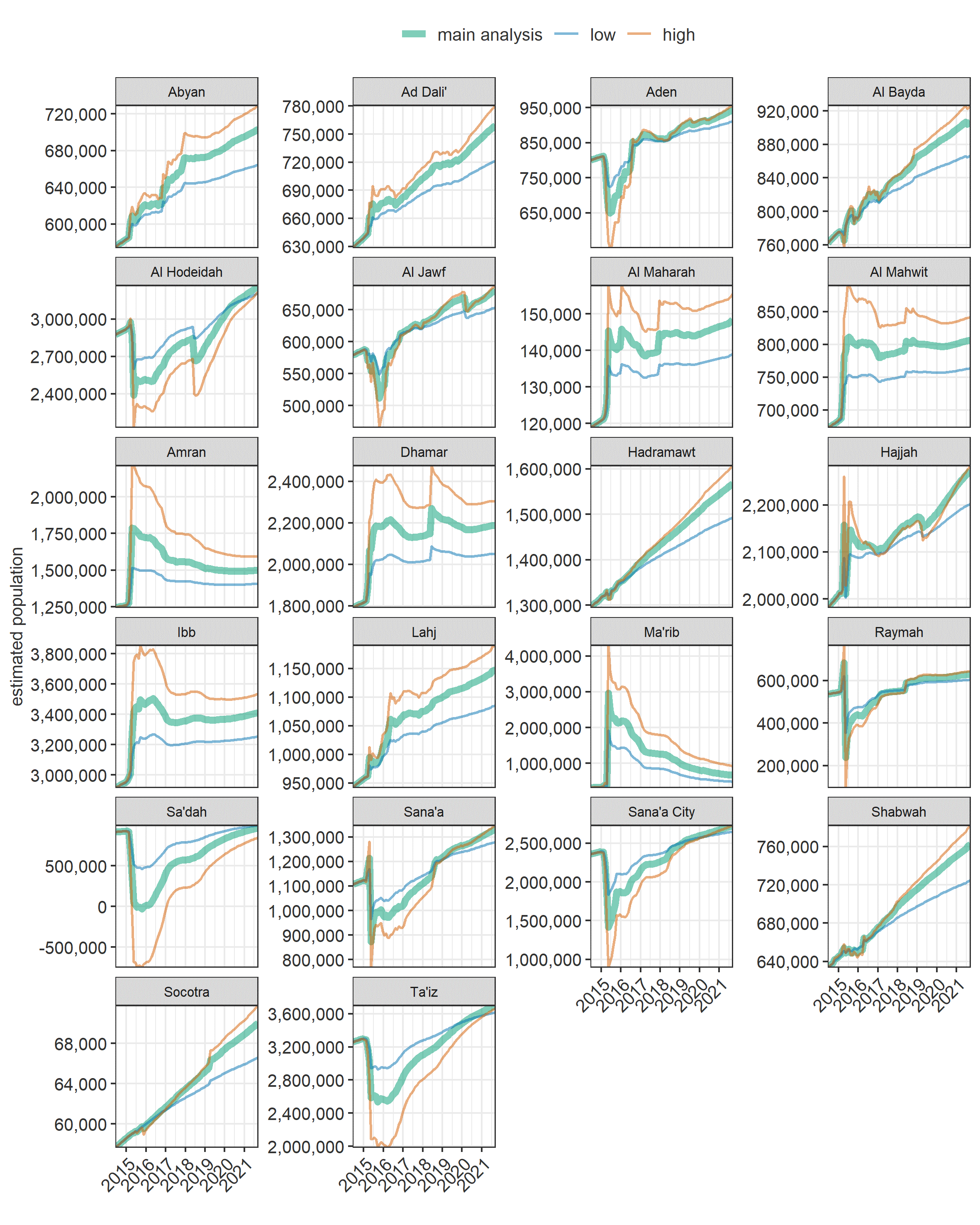

Figure S11. Estimated population of Yemen over time, by governorate and scenario.

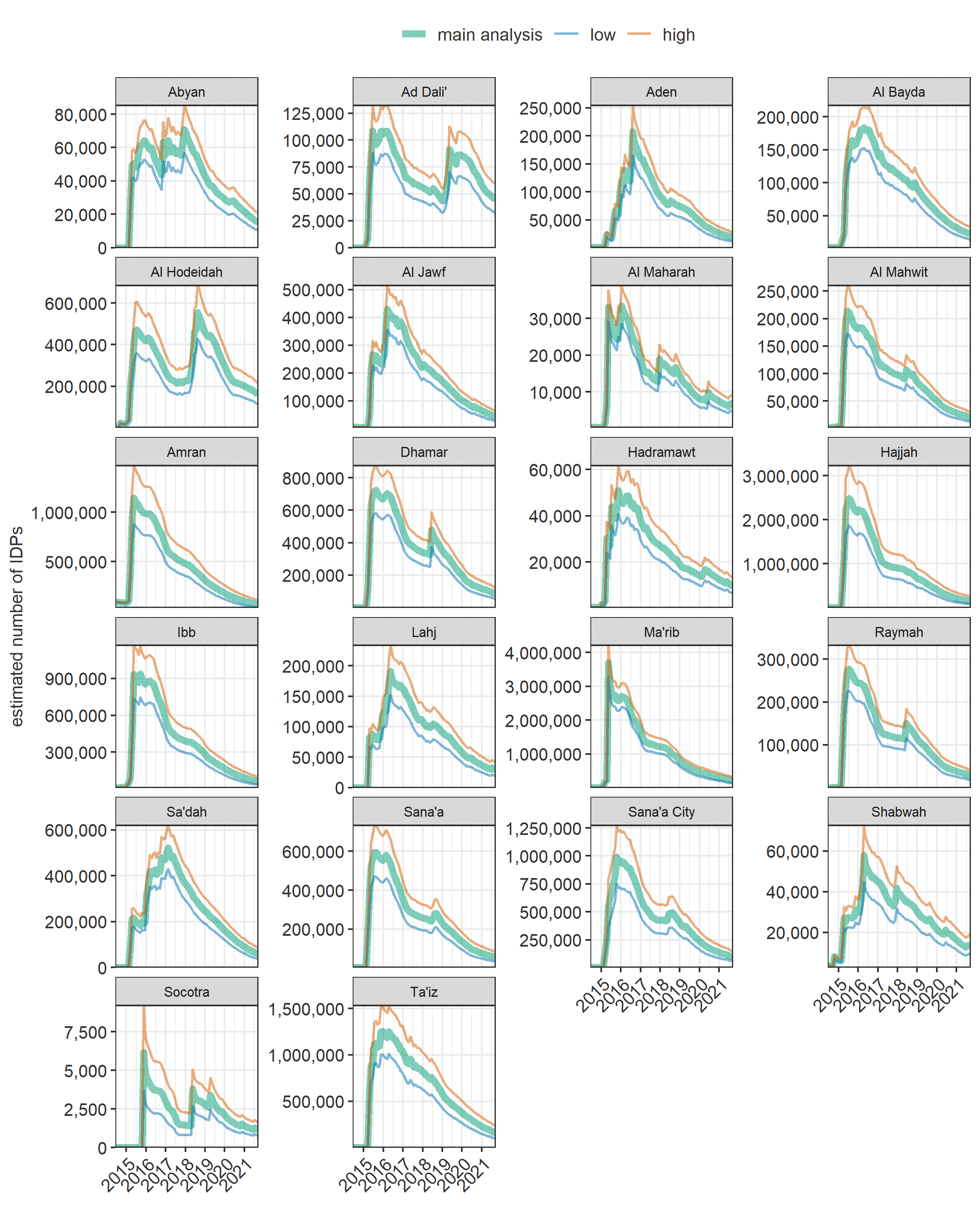

Figure S12. Estimated number of IDPs in Yemen over time, by governorate and scenario.
